# Which curve are we flattening? The disproportionate impact of COVID-19 among economically marginalized communities in Ontario, Canada, was unchanged from wild-type to omicron

**DOI:** 10.1101/2022.10.24.22281104

**Authors:** Huiting Ma, Adrienne K. Chan, Stefan D. Baral, Christine Fahim, Sharon Straus, Beate Sander, Sharmistha Mishra

## Abstract

Economically marginalized communities have faced disproportionately higher risks for infection and death from COVID-19 across Canada. It was anticipated that health disparities would dissipate over time and during subsequent waves. We used person-level surveillance and neighbourhood-level income data to explore, using Lorenz curves and Gini coefficients, magnitude of inequalities in COVID-19 hospitalizations and deaths over five waves of COVID-19 in Ontario, Canada (population 14 million) between February 26, 2020 and February 28, 2022. We found that despite attempts at equity-informed policies alongside fluctuating levels of public health measures, inequalities in hospitalizations and deaths by income remained at levels observed during the first wave - prior to vaccination, discussion or implementation of equity-informed policies - and despite rising levels of hybrid immunity. There was no change in the magnitude of inequalities across all waves evaluated. Our findings indicate that interventions did not sufficiently address differential exposure risks amplified at the intersections of household crowding and size, workplace exposures, and systemic barriers to prevention and care (including access to therapeutics). Equity and effectiveness of programs are inherently linked and ongoing evaluation of both is central to inform the public health response to future waves of COVID-19 and other rapidly emergent pandemics.

## INTRODUCTION

Economically marginalized communities have faced disproportionately higher risks for infection and death from COVID-19 across Canada.^1^ Health disparities were evident early in the pandemic, at the intersections of housing, occupations, and structural racism.^2^ In Ontario, policies attempting to address health inequities were temporarily enacted in subsequent waves, including temporary income support, eviction moratoria, 3-day paid sick leave, and geographic prioritization of vaccine eligibility and allocation. Public health measures designed as restrictions to limit person-to-person contacts continued. It was anticipated that health disparities would be mitigated during subsequent waves. We used metrics of inequality (Lorenz curve and concentration/Gini coefficients) to characterize changes in the magnitude of concentration in COVID-19 hospitalization and deaths by neighbourhood-level income among 14 million Ontario residents across five waves.

## METHODS

We conducted a retrospective, population-based observational study using linked person-level data on all laboratory-confirmed diagnoses of COVID-19 and outcomes in Ontario; the Ontario vaccination database for vaccination status; and Statistics Canada 2016 Census data for dissemination area-level household income per person equivalent; reported using the Strengthening the Reporting of Observational Studies in Epidemiology. The study population comprised individuals who were hospitalized or died following COVID-19 diagnosis between February 26, 2020 and February 28, 2022 (5 waves, **Figure 1**), and excluded travel-related cases and residents of long-term care homes. We generated Lorenz curves for outcomes by DA-level income quintile, and estimated the co-Gini coefficient (zero represents complete equality and one represents complete inequality) in each wave.^3^ Analyses were conducted in R (version 4.0.2). The University of Toronto Health Sciences Research Ethics Board approved the study (protocol #39253).

**Figure 1.**
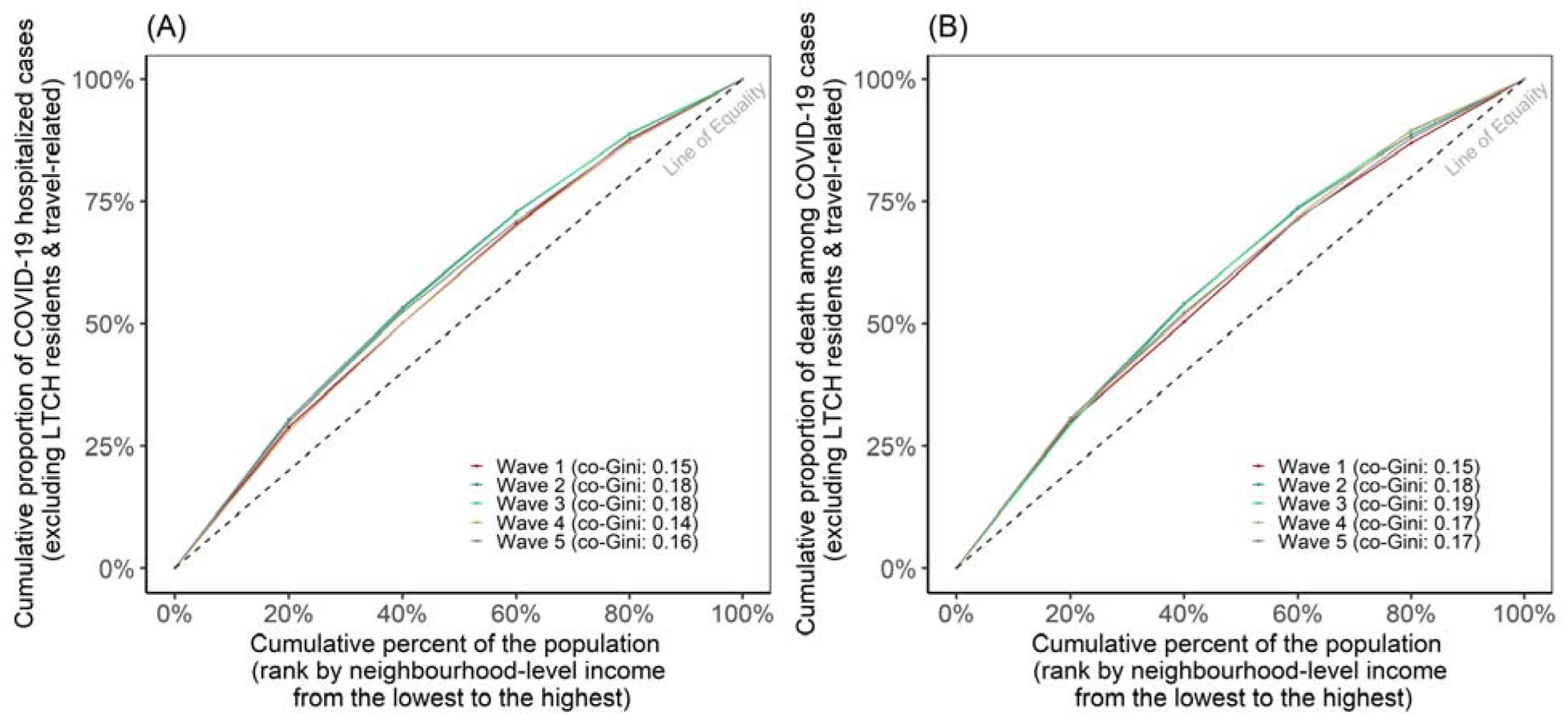
Lorenz curves depicting inequality in COVID-19 hospitalizations (A) and deaths (B) by income for each of the five waves. Wave 1 (wild-type): February 26, 2020 to August 31, 2020 (length of wave 188 days); Wave 2 (wild-type and emergence of alpha): September 1, 2020 to February 28, 2021 (length of wave 181 days); Wave 3 (alpha and emergence of delta): March 1, 2021 to July 31, 2021 (length of wave 153 days); Wave 4 (delta): August 1, 2021 to December 14, 2021 (length of wave 135 days); Wave 5 (predominantly omicron): December 15, 2021 to February 28, 2022 (length of wave 75 days).^6^ The cumulative number of hospitalizations and deaths due to COVID-19 in each wave were: Wave 1: 3611, 935; Wave 2: 9590, 2295; Wave 3: 11901, 2184; Wave 4: 3475, 531; Wave 5: 9425, 2143. The co-Gini coefficient refers to the magnitude of concentration or inequality. Neighbourhood-level income refers to the per-person equivalent, after-tax income and accounting for regional cost of living; and is derived from the 2016 Census. The further the Lorenz curve from the line of equality, the greater the inequality/concentration.

## RESULTS

Over the study period, hospitalizations and deaths were concentrated among the 20% of the population living in the lowest income neighbourhoods (**Figure 1, Figure S1**). The magnitude of inequality in hospitalization and mortality by income remained unchanged across waves (range in co-Gini, 0.14-0.19), despite less inequality by income in vaccination or known past infection (**Figure 2**). Vaccine coverage was consistently higher in high-income areas (especially with third doses, **Figure S2-3**), while rates of prior infection were higher in lower-income areas (**Figure S1**). Together, the magnitude of inequality in hospitalizations and deaths were 10-fold higher than magnitude of inequality in vaccination/past infection (**Figures 1-2, Figure S1 and Figure S4**).

**Figure 2.**
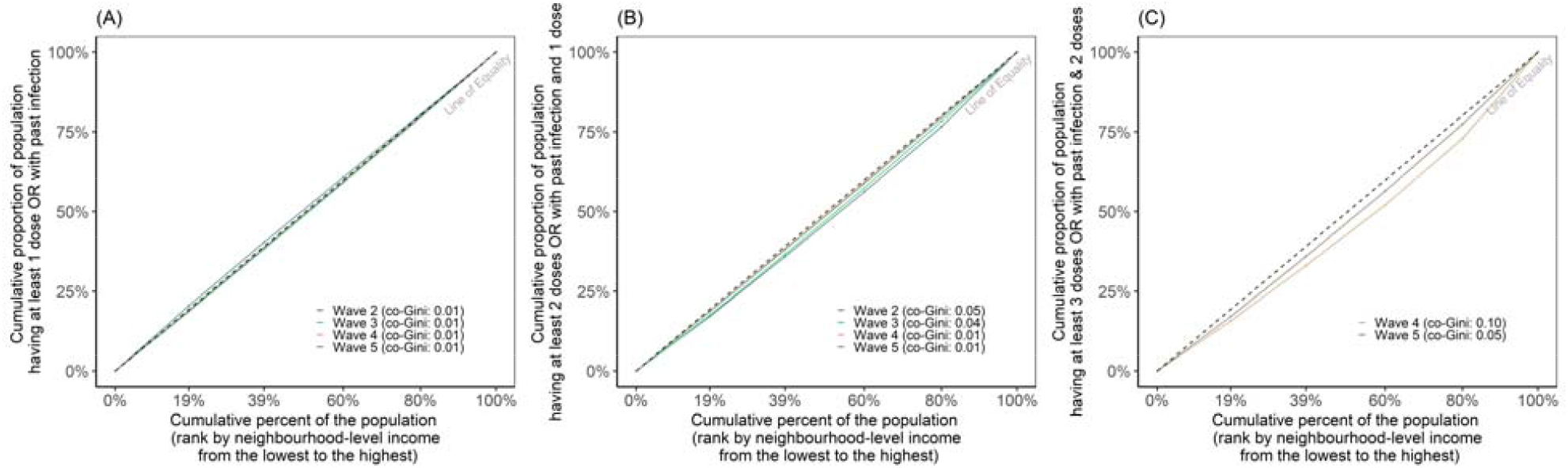
Lorenz curves depicting level of vaccination and/or past infection by income within each of the five waves. Panel A, B, and C respectively depict: proportion with one vaccine dose or a prior diagnosis; proportion with a 2nd dose or a first dose with a prior diagnosis; proportion with a third dose or two doses with a prior diagnosis. Wave 1 (wild-type): February 26, 2020 to August 31, 2020 (length of wave 188 days); Wave 2 (wild-type and emergence of alpha): September 1, 2020 to February 28, 2021 (length of wave 181 days); Wave 3 (alpha and emergence of delta): March 1, 2021 to July 31, 2021 (length of wave 153 days); Wave 4 (delta): August 1, 2021 to December 14, 2021 (length of wave 135 days); Wave 5 (predominantly omicron): December 15, 2021 to February 28, 2022 (length of wave 75 days).^6^ Neighbourhood-level income refers to the per-person equivalent, after-tax income and accounting for regional cost of living; and is derived from the 2016 Census. The further the Lorenz curve from the line of equality, the greater the inequality/concentration. The denominator is Ministry of Heath population estimates in 2021 and the numerator is the cumulative vaccine number + cumulative infection number - cumulative reinfection number (regardless when infection occurred).

## DISCUSSION

Despite attempts at equity-informed policies alongside fluctuating levels of public health measures, the magnitude of inequalities in hospitalizations and deaths by income remained at levels observed during the first wave - prior to vaccination, and discussion or implementation of equity-informed policies - and despite rising levels of hybrid immunity.

Our analysis follows a descriptive metric of inequality over time signaling the need for future explanatory modeling to explore mechanisms that perpetuated health disparities to inform responses. Comorbidities, for example, are confounders, but prior studies consistently demonstrated that social determinants remain a critical determinant of COVID-19 deaths at a population-level even after accounting for comorbidities.^4^ The small magnitude of inequality in past infection and vaccination suggest differential levels of hybrid immunity alone may be insufficient to explain our findings.

Our findings indicate that interventions did not sufficiently address differential exposure risks amplified at the intersections of household crowding and size, workplace exposures, and systemic barriers to prevention and care (including access to therapeutics). Equality in intervention reach is often insufficient to redress longstanding inequities and overall reductions in cases and deaths are insufficient metrics of comparative success^5^ if existing health disparities persist. Equity and effectiveness of programs are inherently linked–and ongoing evaluation of both is central to inform the public health response to future waves of COVID-19 and other rapidly emergent pandemics.

## Data Availability

Reported COVID-19 cases were obtained from the Case and Contact Management Solutions and vaccination status were obtained from the provincial COVaxON Vaccination Management System via the Ontario COVID-19 Modelling Consensus Table and with approval from the University of Toronto Health Sciences Research Ethics Board (protocol no. 39253). The analyses, conclusions, opinions and statements expressed herein are solely those of the authors and do not reflect those of the funding or data sources; no endorsement is intended or should be inferred.

## ACKNOWLEDGEMENTS

We thank the Ontario Ministry of Health Data Analytics Branch for supporting access to data via the Ontario COVID-19 Modelling Consensus Table. SM is supported by a Tier 2 Canada Research Chair in Mathematical Modeling and Program Science. BS is supported by a Tier 2 Canada Research Chair in Economics of Infectious Diseases. SS is supported by a Tier 1 Canada Research Chair in Knowledge Translation and Quality of Care.

## AUTHOR CONTRIBUTIONS

HM, AC, and SM conceived of and designed the study. HM, AC, and SM conducted literature review. HM developed the analysis plan with input from AC and SM. HM executed the analysis plan and conducted the statistical analysis. HM and AC wrote the first draft of manuscript. All authors (HM, AC, SB, CF, SS, BS, and SM) provided critical input into study design, interpretation of results, and manuscript review and editing.

## DATA STATEMENT

Data statement: Reported COVID-19 cases were obtained from the Case and Contact Management Solutions and vaccination status were obtained from the provincial COVaxON Vaccination Management System via the Ontario COVID-19 Modelling Consensus Table and with approval from the University of Toronto Health Sciences Research Ethics Board (protocol no. 39253). The analyses, conclusions, opinions and statements expressed herein are solely those of the authors and do not reflect those of the funding or data sources; no endorsement is intended or should be inferred.

