## Supplemental Materials for "Which curve are we flattening? The disproportionate impact of COVID-19 among economically marginalized communities in Ontario, Canada, was unchanged from wild-type to omicron"

**Supplemental Appendix**

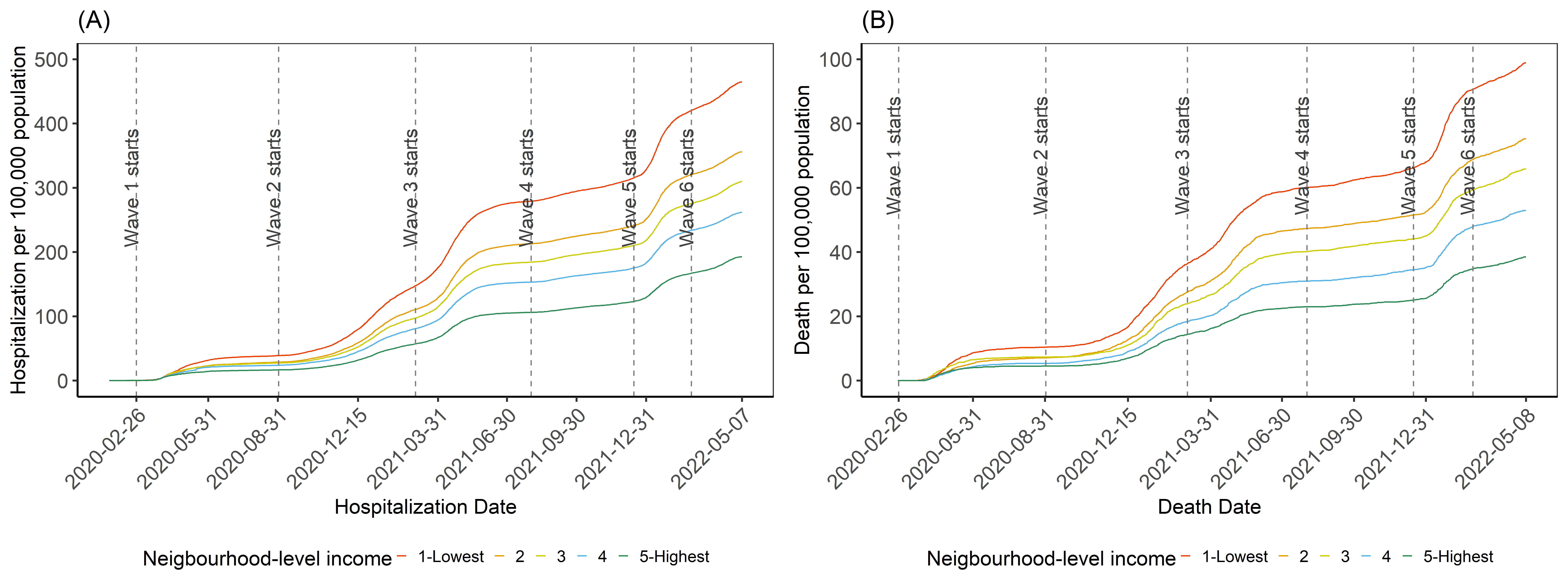

**Figure S1. Cumulative per-capita COVID-19 hospitalization (A) and Death (B) over time by income.** Wave 1 (wild-type): February 26, 2020 to August 31, 2020 (length of wave 188 days); Wave 2 (wild-type and emergence of alpha): September 1, 2020 to February 28, 2021 (length of wave 181 days); Wave 3 (alpha and emergence of delta): March 1, 2021 to July 31, 2021 (length of wave 153 days); Wave 4 (delta): August 1, 2021 to December 14, 2021 (length of wave 135 days); Wave 5 (predominantly omicron): December 15, 2021 to February 28, 2022 (length of wave 75 days).^6^ Neighbourhood-level income (after-tax, per-person equivalent) quintiles where the lowest quintile (quintile 1) represents areas with the lowest average total after-tax income and the highest quintile (quintile 5) represents areas with the highest average total after-tax income.

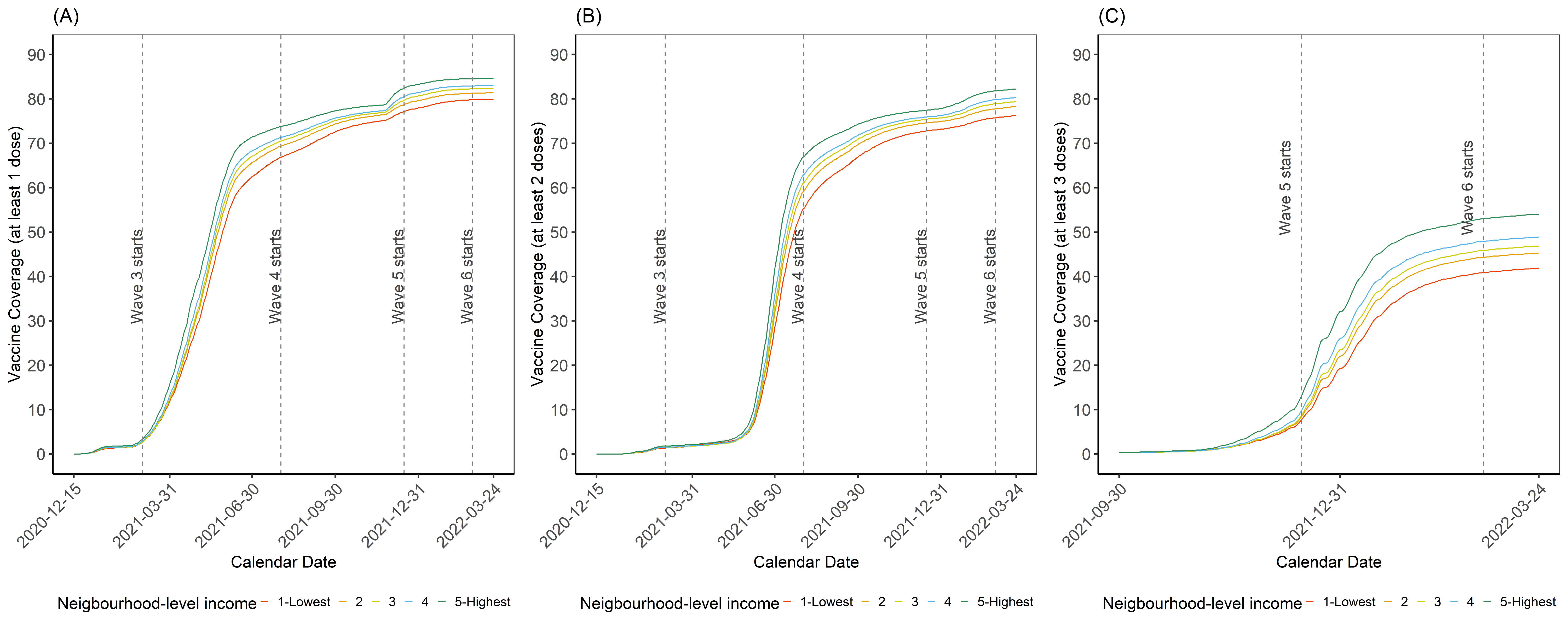

**Figure S2. Vaccination coverage of at least 1 dose (A), at least 2 doses (B) and at least 3 doses (C) over time by income.** Wave 1 (wild-type): February 26, 2020 to August 31, 2020 (length of wave 188 days); Wave 2 (wild-type and emergence of alpha): September 1, 2020 to February 28, 2021 (length of wave 181 days); Wave 3 (alpha and emergence of delta): March 1, 2021 to July 31, 2021 (length of wave 153 days); Wave 4 (delta): August 1, 2021 to December 14, 2021 (length of wave 135 days); Wave 5 (predominantly omicron): December 15, 2021 to February 28, 2022 (length of wave 75 days).^6^ Neighbourhood-level income (after-tax, per-person equivalent) quintiles where the lowest quintile (quintile 1) represents areas with the lowest average total after-tax income and the highest quintile (quintile 5) represents areas with the highest average total after-tax income.

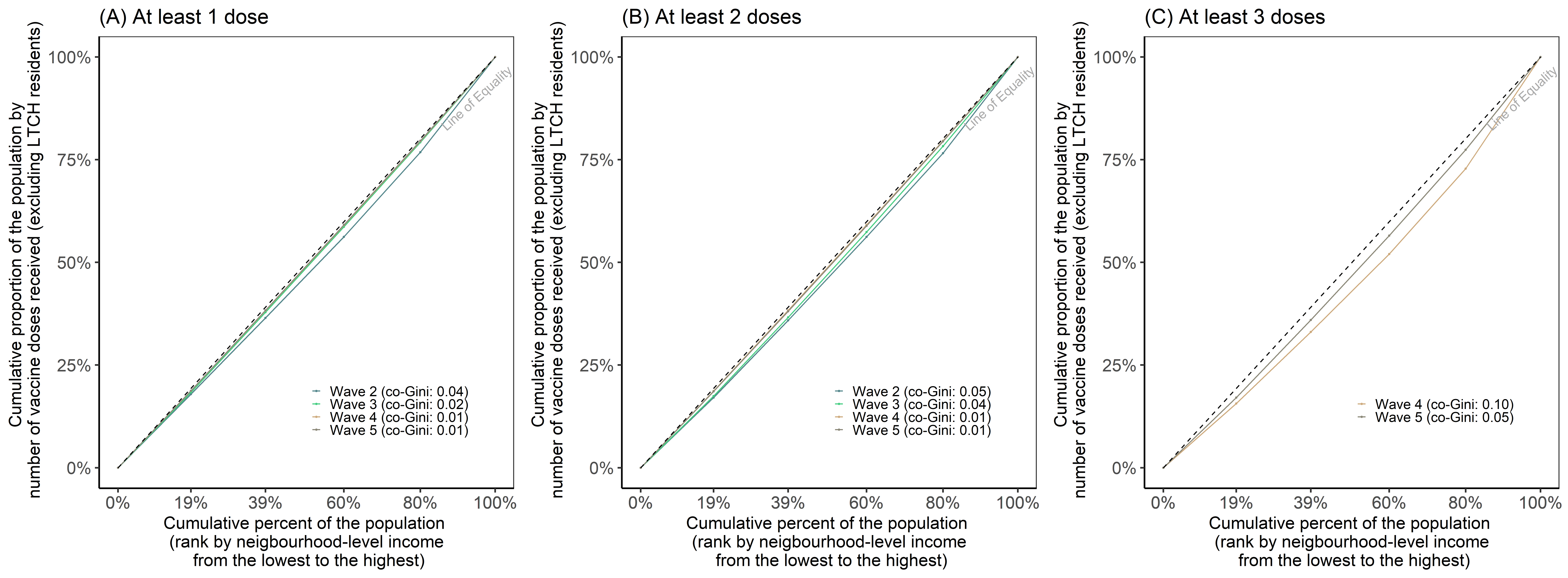

**Figure S3. Lorenz curves of at least 1 dose (A), at least 2 doses (B), and at least 3 doses (C) of vaccination by income.** Wave 1 (wild-type): February 26, 2020 to August 31, 2020 (length of wave 188 days); Wave 2 (wild-type and emergence of alpha): September 1, 2020 to February 28, 2021 (length of wave 181 days); Wave 3 (alpha and emergence of delta): March 1, 2021 to July 31, 2021 (length of wave 153 days); Wave 4 (delta): August 1, 2021 to December 14, 2021 (length of wave 135 days); Wave 5 (predominantly omicron): December 15, 2021 to February 28, 2022 (length of wave 75 days).^6^ Neighbourhood-level income (after-tax, per-person equivalent) quintiles where the lowest quintile (quintile 1) represents areas with the lowest average total after-tax income and the highest quintile (quintile 5) represents areas with the highest average total after-tax income.

**Figure S4. Cumulative proportion over time of population by vaccine and known prior infection, by income.** Panel A, B, and C respectively depict: proportion with one vaccine dose or a prior diagnosis; proportion with a 2nd dose or a first dose with a prior diagnosis; proportion with a third dose or two doses with a prior diagnosis. Wave 1 (wild-type): February 26, 2020 to August 31, 2020 (length of wave 188 days); Wave 2 (wild-type and emergence of alpha): September 1, 2020 to February 28, 2021 (length of wave 181 days); Wave 3 (alpha and emergence of delta): March 1, 2021 to July 31, 2021 (length of wave 153 days); Wave 4 (delta): August 1, 2021 to December 14, 2021 (length of wave 135 days); Wave 5 (predominantly omicron): December 15, 2021 to February 28, 2022 (length of wave 75 days).^6^ Neighbourhood-level income (after-tax, per-person equivalent) quintiles where the lowest quintile (quintile 1) represents areas with the lowest average total after-tax income and the highest quintile (quintile 5) represents areas with the highest average total after-tax income.

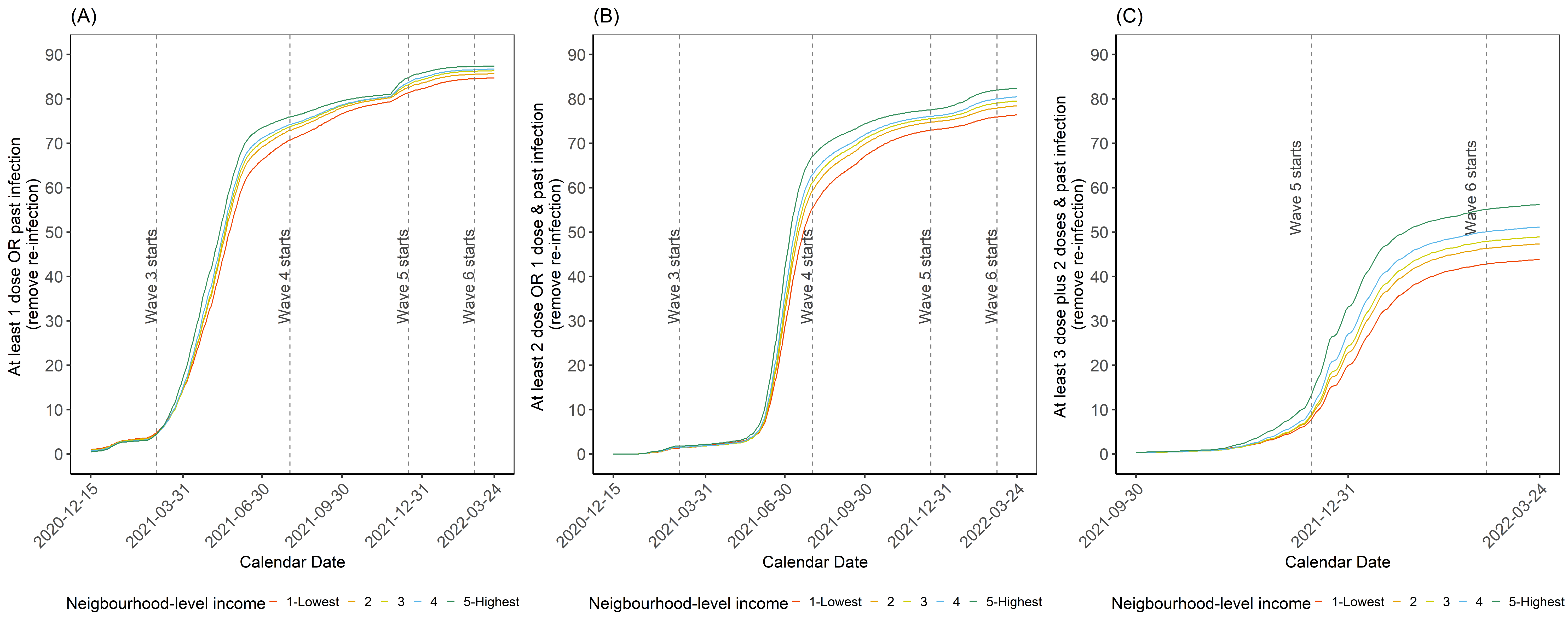
